# Evaluation of effects of public health interventions on COVID-19 transmission for Pakistan: A mathematical simulation study

**DOI:** 10.1101/2020.04.30.20086447

**Authors:** Zulfiqar A Bhutta, Ofir Harari, Jay JH Park, Noor-E Zannat, Michael Zoratti, Tim Churches, Kristian Thorlund, Edward J Mills

## Abstract

**Background:** In an effort to contain the COVID-19 epidemic, many governments across the world have enforced lockdown or social distancing measures. Several outbreak models have been developed to investigate the effects of different public health strategies for COVID-19, but they have not been developed for Pakistan and other South East Asian countries, where a large proportion of global population resides.

**Methods:** We developed a stochastic individual contact model by extending the widely-used Susceptible-Infectious-Recovered (SIR) compartment model with additional compartments to model both anticipated mitigating effects of public health intervention strategies for Pakistan. We estimated the projected spread, number of hospitalizations, and case fatalities under no intervention and four increasingly stringent public health strategies of social distancing and self-isolation at the national and provincial levels of Pakistan.

**Results:** Our analysis shows that without any public health interventions the expected number of cumulative case fatalities is 671,596 in Pakistan with the virus is expected to peak in terms of the number of required ICU-hospitalizations at 198,593 persons by the end of the June 2020. The estimated total numbers of cumulative case fatalities are lower for other public health strategies with strict social distancing showing the lowest number of deaths at 1,588 (Self-isolation: n=341,359; Flexible social distancing strategy: n=3,995; and Exit strategy: n=28,214). The lowest number of required ICU-hospitalization is also estimated for strict social distancing strategy (n=266 persons at the end of May 2020). Generally, the simulated effects of the different public health strategies at the provincial-level were similar to the national-level with strict social distancing showing the fewest number of case fatalities and ICU-hospitalizations.

**Conclusion:** Our results indicate that case fatalities and ICU-hospitalizations for Pakistan will be high without any public health interventions. While strict social distancing can potentially prevent a large number of deaths and ICU-hospitalizations, the government faces an important dilemma of potentially severe economic downfall. Consideration of a temporary strict social distancing strategy with gradual return of the lower-risk Pakistani population, as simulated in our exit strategy scenario, may an effective compromise between public health and economy of Pakistani population.

## Introduction

On January 30, 2020, the World Health Organization (WHO) formally declared the outbreak of novel coronavirus a Global Public Health Emergency of International Concern.^1^ Within two months of that declaration, the SARS-CoV-2 virus has led to the first global pandemic in over a century. The numbers of confirmed cases and COVID-19 related deaths continue to rise at a rapid rate, with major effects on local and the global economy, and with no clear end in sight.^2^ In an effort to contain the epidemic, governments in many countries have enforced lock-downs and other social distancing measures in various forms.

Over the past few months, several national and provincial disease outbreak models have been developed to investigate the likely effects of strategies for slowing or mitigating the spread of COVID-19.^3–10^ As the pandemic spread to European countries and North America in late February, several models have started to focus on these specific geographic regions.^11–13^ Since the pandemic is now emerging in other regions, these is an increasing need to modify existing models to the settings pertinent to other resource-limited geographic regions such as South East Asian countries, where a large proportion of global population resides.

In this paper, we present a stochastic individual contact model (ICM) based on an extension of the widely-used Susceptible-Infectious-Recovered (SIR) compartment differential equation model^14,15^ to simulate the COVID-19 outbreak in Pakistan under different public health interventions and to advise the Government of Pakistan and public health authorities on their policy response to COVID-19. In this study, we estimated the projected spread, number of hospitalizations, and case fatalities under no intervention and four increasingly stringent public health strategies of social distancing and self-isolation at the national and provincial levels of Pakistan.

## Methods

### Model structure

To evaluate the effects of public health interventions on COVID-19 and forecast its spread in Pakistan, we conducted a simulation study using a computational stochastic individual contact model (ICM) based on an extension of the *Susceptible-Infectious-Recovered* (SIR) compartment model.^15,16^ This model comprises seven compartments as illustrated in Figure 1 (see Supplementary Table 1 for further details). Three components are similar to SIR compartment model: The S compartment denotes susceptible individuals; the I compartment denotes symptomatic individuals who are both infected with COVID-19 and infectious to others; and the R compartment denotes individuals who have recovered from COVID-19 and are no longer infectious. The SIR model was expanded with the addition of four compartments (E, Q, H, and F) to model both anticipated mitigating effects of public health intervention strategies as well as measurable impact on public health. Unlike the E compartment in traditional SEIR models, the E compartment in our model denotes asymptomatic COVID-19-positive individuals who are infectious, in order to enable simulation of transmission during the COVID-19 incubation period, as reported by several investigators^17^; the Q compartment represents symptomatic (or test-positive) infectious individuals who are self-isolating or in supervised isolation; the H compartment represents individuals who require hospitalization (if the number of required hospitalizations is below the hospital capacity, then it is assumed in the model that these individuals would be hospitalized, but if hospital capacity is exceeded then the excess portion of those requiring hospitalisation remain not hospitalised, with consequently higher mortality for that fraction of cases); and the F compartment denotes case fatalities due to COVID-19.

**Figure 1:**
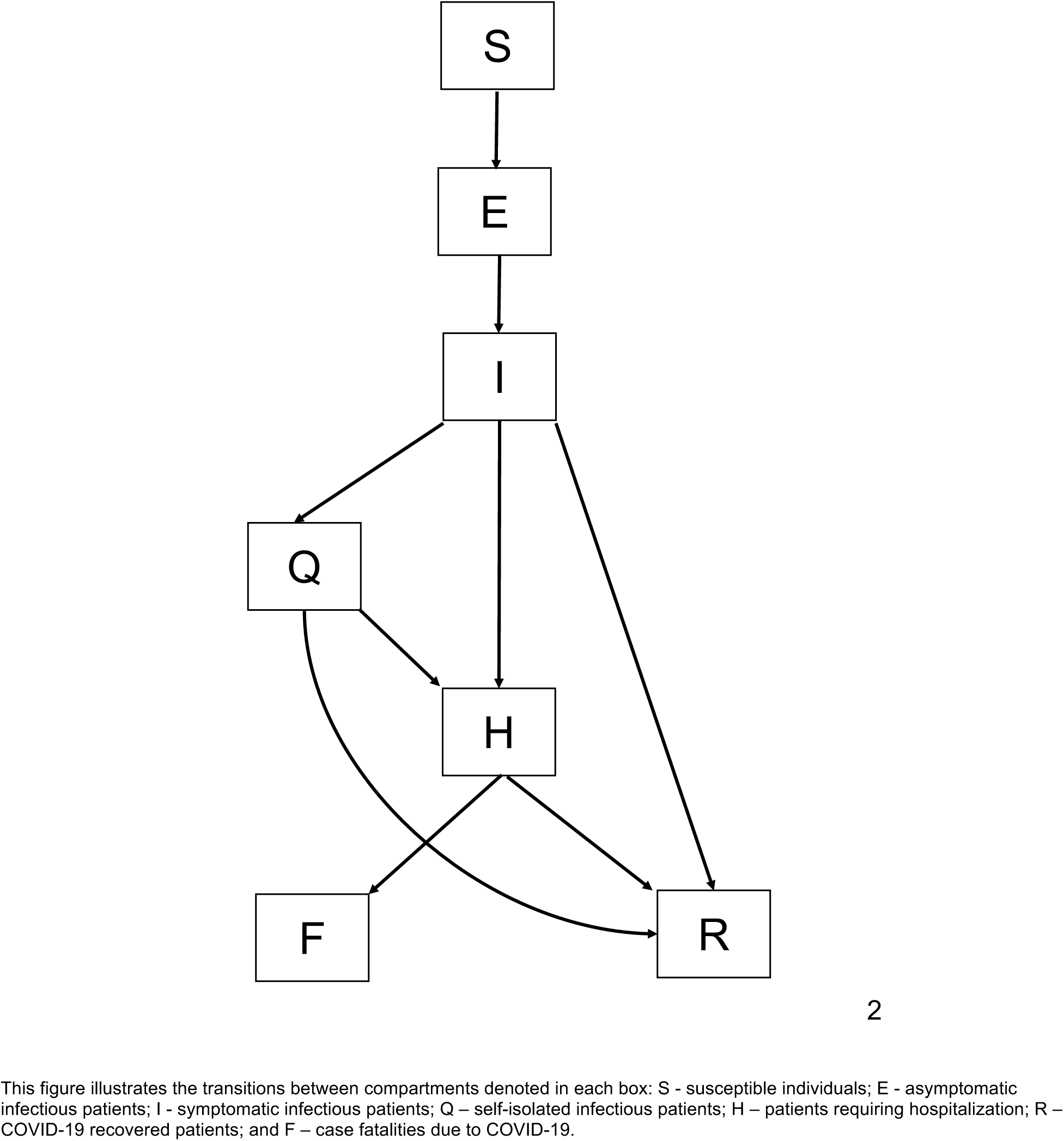
Transition diagram of this compartment model.

### Model parameters

Model parameters were populated using national-level datasets from Pakistan and other empirical estimates (Supplementary Table 2). The infectivity between infectious and susceptible individuals was calculated based on the daily average number of exposures between susceptible individuals and infectious individuals with the probability of passing on infection at each exposure event between asymptomatic and symptomatic individuals with the susceptible individuals. In this approach, the daily average number of exposures between susceptible individuals (S) and infectious individuals on average was assumed to have 10 exposures per day (per infected individual) at the national-level. This number was determined under the assumption that, with no intervention, 75% of the population would eventually become infected. The probability of passing on infection at each of the exposure event was assumed to be 2.4% between symptomatic and susceptible individuals and 1.6% between asymptomatic and susceptible individuals, representing presumed lower average viral shedding by infected but asymptomic individuals. Given 10 exposures per day assumed, the probability of 2.4% infectivity for the symptomatic individuals translates to an average symptomatic individual infecting an average of 2.4 individuals over a 10-day span, which in turn is cognate to the basic reproduction number used to parameterise infectivity in traditional mathematical models.

For the projections at the provincial level, we used the reported number of cases for each province at the starting date of the simulation for the “Q” compartment and made the assumption that the number of asymptomatic (E) and symptomatic (I) cases in the community equalled it. We also took the different population sizes and areas of each province into account. The average number of daily exposures per infected individual at the provincial level was determined, assuming a national level of 10 exposures per day and the daily exposure rate in urban areas would be three times higher than the rural areas, while considering the population distribution of urban and rural areas for each province (Supplementary Table 3). Provincial level estimates on the number of hospital beds were used as the hospital capacity for the model.^18^

### Intervention scenarios

We considered four different public health intervention scenarios (Table 1). *Scenario 1* was based on a ‘no intervention’ approach, where the disease would run its natural course until the rates of recovery would surpass the rates of new infections, resulting in increasing levels of “herd immunity” at a national level. Scenario 2 was based on ‘self-isolation’, and we assumed that just 3% of symptomatic cases would self-isolate each day at baseline. However, starting at the 15^th^ day of the simulation, mimicking a combination of increased testing and raising of awareness in the community, the proportion of symptomatic individuals who would self-isolate (or be placed in supervised isolation) each day would linearly increase from 3% to 25% over the course of 15 days. The same self-isolation regime was assumed for social distancing (scenarios 3 and 4). Scenario 3 was based on ‘strict social distancing’. It assumed the same self-isolation as scenario 2 (i.e., days 15 to 30), but in addition, imposes strict restrictions on public access from the 30^th^ day to the extent that the average number of daily exposures of infectious individuals to susceptible individuals will decrease from an initial 10 (see model parameters) to 2.5 over the course of the day 30 to day 45. In dense urban areas, this scenario is analogous to a government lockdown scenario. Scenario 4 was based on ‘flexible social distancing’, where day 15 to 30 are identical to scenario 2, but where in addition the number of daily exposures gradually decreases from 10 to 5 over the course of subsequent 15 days. Scenario 5 is referred to as ‘exit strategy’ scenario where after 30 days of lock-down (as in scenario 3), the exposure rate gradually increases until it reaches 70% of the original rate, reflecting 15% vulnerable individuals remaining in a lock-down along with their caregivers. During the exit period, the hospitalization rate starts at 70% of the original value at the start of the simulation and gradually increases to 85%.

**Table 1:** Summary of public health interventions considered.

| Scenarios | Self-isolation of COVID-19 positive | Social distancing | Modeling start date* | Social intervention start date |
| --- | --- | --- | --- | --- |
| 1. No intervention | No | No | April 5<sup>th</sup>, 2020 | -- |
| 2. Self-isolation | Yes | No | April 5<sup>th</sup>, 2020 | April 8<sup>th</sup>, 2020 |
| 3. Strict social distancing | Yes | High compliance | April 5<sup>th</sup>, 2020 | April 20<sup>th</sup>, 2020 |
| 4. Flexible social distancing | Yes | Moderate compliance | April 5<sup>th</sup>, 2020 | April 20<sup>th</sup>, 2020 |
| 5. Exit strategy (after strict social distancing) | Yes | High compliance for one month and gradual return* | April 5<sup>th</sup>, 2020 | April 20<sup>th</sup>, 2020 |

For all scenarios, the modeling start date was defined to be April 5<sup>th</sup>, 2020. The first date modelled has started with 1,000 infected and quarantined, 1,000 infected and not-quarantined, and 1,000 asymptomatic cases. The modelled scenarios and dates take into account for the time taken for the impact and scale-up of self-isolation and social distancing strategies in Pakistan and the variation of population size and urban and rural distribution between different provinces.
\*The exit strategy intervention scenario starts with strict social distancing for one month then gradual return of the general population other than high risk population and their caregivers

### Analysis

Using the base values, we estimated the number of individuals for each of the seven compartments on a weekly basis. Each model run began on April 5^th^, 2020 and simulated the spread of the disease in the population for one calendar year. For each province, 1000 simulation iterations were run to improve the reliability of the predictions, given the stochastic nature of the model. We recorded the number of individuals in each compartment each day as well as the number of hospitalizations – of which 15% were assumed to require critical care – and case fatalities. Evolution of the pandemic was assessed graphically by plotting the number of individuals in each compartment against time, as well as the number of hospitalizations and case fatalities against time. Base case scenario input parameters and their justifications are presented in Supplementary Table 2.

We also conducted deterministic sensitivity analyses using ‘worst’ and ‘best’ cases on parameters related to infectivity of asymptomatic patients and the mortality rate among those who require hospitalization (Supplementary Table 4). For the infectivity of asymptomatic patients, we assumed that they would have the same infectivity (2.4%) as the symptomatic patients as the worst-case scenario (the base case assumed 1.6% for the asymptomatic patients), and for the best-case, we assumed one-third infectivity (0.8%) for the symptomatic patients. For baseline mortality rate among those requiring hospitalization, we assumed that baseline daily mortality rate for people needing hospitalization would be 50% higher as the worst case; for the best cases, we assumed that the daily mortality rate for those requiring hospitalization would be 50% lower than the base case. There were three sensitivity scenario analyses: the first scenario used different asymptomatic infectivity assumption alone; the second scenario assumed different baseline mortality rates, and the third scenario used combination of worst-case and base-case of asymptomatic infectivity and baseline mortality rate.

## Results

### Base case

Figure 2 illustrates the national-level estimates of cumulative case fatalities from May 1^st^, 2020 to April 30^th^, 2021 (sensitivity bands of fatalities rates also shown). Based on the simulations, the absence of public health interventions (Scenario 1: No intervention) results in the highest numbers of case fatalities (Table 2). Under this scenario, total expected number of cumulative case fatalities is 671,596. While the self-isolation strategy (Scenario 2) yielded approximately half as many case fatalities as the no intervention strategy, this showed the second highest number of cumulative case fatalities at 341,359. The strict social distancing strategy (Scenario 3) showed the fewest total number of cumulative case fatalities estimated at 1,588. With flexible social distancing (Scenario 4), the estimated total number of cumulative case fatalities is higher estimated at 3,995. Under the exit strategy (Scenario 5), the total number of cumulative case fatalities is estimated at 28,214 by March 2021.

**Figure 2:**
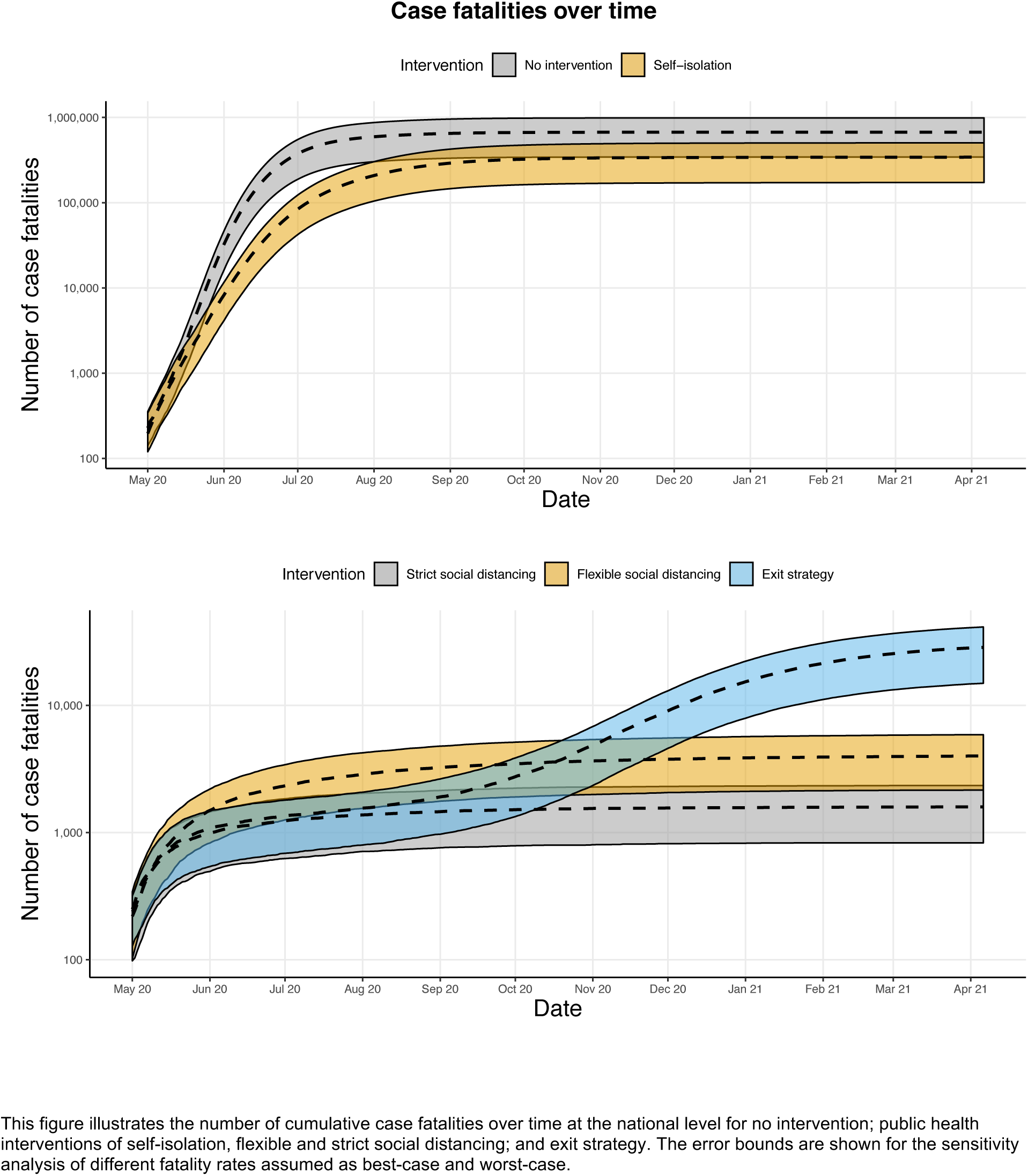
National-level estimates of cumulative case fatalities spread with sensitivity bands of fatality rates.

**Table 2:** National-level estimates of ICU-hospitalization and cumulative case fatalities between no intervention and different public health interventions.

| Time | Case fatalities |  |  |  |  | ICU-hospitalization |  |  |  |  |
| --- | --- | --- | --- | --- | --- | --- | --- | --- | --- | --- |
|  | No intervention | Self-isolation | Flexible social distancing | Strict social distancing | Exit strategy | No intervention | Self-isolation | Flexible social distancing | Strict social distancing | Exit strategy |
| May 2020 | 28,153 | 7,492 | 1,462 | 975 | 1,062 | 75,397 | 16,819 | 903 | 266 | 268 |
| June 2020 | 362,884 | 79,930 | 2,306 | 1,234 | 1,342 | 198,593 | 66,306 | 575 | 94 | 114 |
| July 2020 | 590,577 | 204,811 | 2,856 | 1,372 | 1,547 | 50,760 | 59,856 | 368 | 52 | 200 |
| Aug 2020 | 649,086 | 289,509 | 3,236 | 1,455 | 1,889 | 16,950 | 31,609 | 248 | 38 | 490 |
| Sept 2020 | 666,327 | 323,011 | 3,460 | 1,508 | 2,700 | 4,715 | 12,027 | 184 | 22 | 1,181 |
| Oct 2020 | 670,510 | 334,649 | 3,650 | 1,546 | 4,764 | 972 | 4,358 | 112 | 15 | 2,618 |
| Nov 2020 | 671,333 | 338,357 | 3,769 | 1,558 | 8,879 | 223 | 1,679 | 80 | 9 | 4,233 |
| Dec 2020 | 671,535 | 339,844 | 3,860 | 1,568 | 15,100 | 61 | 955 | 60 | 3 | 4,830 |
| Jan 2021 | 671,582 | 340,783 | 3,919 | 1,579 | 21,222 | 12 | 638 | 41 | 2 | 4,235 |
| Feb 2021 | 671,594 | 341,359 | 3,957 | 1,586 | 25,380 | 2 | 497 | 30 | 1 | 3,041 |
| March 2021 | 671,596 | 341,861 | 3,995 | 1,588 | 28,214 | 2 | 422 | 23 | 2 | 1,785 |

Numbers shown indicates the number of case fatalities and ICU-hospitalization required at the end of each month at the national level for no intervention; public health interventions of self-isolation, flexible and strict social distancing; and exit strategy.

The temporal trends of national-level estimates of ICU-hospitalization requirements over are shown in the Appendix (Supplementary Figures 1 and 2). Without any public health intervention, the COVID-19 virus is expected to peak in terms of the number of required ICU-hospitalizations at 198,593 persons by the end of the June 2020 (Table 2). With self-isolation, the COVID-19 virus, on the other hand, is expected to peak in June 2020 with 66,306 patients requiring ICU hospitalization for COVID-19. With the strict social distancing strategy, the COVID-19 virus is expected to peak in terms of the number of required ICU-hospitalizations at 266 persons by the end of May 2020. With flexible social distancing, the highest number of required ICU hospitalizations of 903 is expected around May 2020. Under the exit strategy scenario, the COVID-19 virus is expected to peak in terms of the number of required ICU-hospitalizations at 4,830 persons by the end of December 2020, that could be potentially manageable by the local health system at capacity.

The base-case results of cumulative case fatalities and ICU-hospitalization requirements are provided in the Appendix for each of the five major provinces in Pakistan: Balochistan (Supplementary Table 5); Islamabad Capital Territory (Supplementary Table 6); Khyber Pakhtunkhwa and Federally Administered Tribal Area (KP-FATA; Supplementary Table 7); Punjab (Supplementary Table 8); and Sindh (Supplementary Table 9). Generally, the simulated effects of the different public health intervention strategies at the provincial-level were similar to the national-level with no intervention strategy showing the highest number of case fatalities and ICU-hospitalizations at each of the provinces and strict social distancing strategy showing the fewest number of case fatalities and ICU-hospitalizations. Without any public health intervention, Punjab where 53% of the Pakistani population reside according to the 2017 census, the highest number of case fatalities (n = 370,050) and required ICU-hospitalizations (n = 148,327 in June 2020) were observed from our simulations. The province of Balochistan that has the lowest urban-to-rural ratios, on the other hand, showed the fewest number of case fatalities (n=36,188) and required-ICU hospitalizations (n = 10,285 in June 2020) without any public health intervention strategy.

### Sensitivity analyses

Figure 3 and Figure 4 illustrate the national-level estimates of cumulative case fatalities during the same period with sensitivity bands of different case fatality rates and different infectivity of asymptomatic patients, respectively. The sensitivity analysis results on the ICU-hospitalization requirements with respect of case fatality rates are shown in Supplementary Figure 3, and the sensitivity analyses in terms of infectivity of asymptomatic patients are shown in Supplementary Figure 4. Modifications to the parameter of infectivity of asymptomatic patients returned similar trends as observed under the base case scenarios. Again, consistent with the case analyses, the scenario of no public health intervention resulted in the worst outcomes with respect to the number of case fatalities and number of persons requiring hospitalization. Similarly, the strict social distancing strategy showed the smallest number of case fatalities and number of persons requiring hospitalization. Sensitivity analyses simultaneously evaluating changes in parameters of infectivity and mortality rates showed similar trends to both the base case analysis and other sensitivity analyses at the national (Supplementary Tables 10 and 11) and provincial levels (Supplementary Tables 12 and 13).

**Figure 3:**
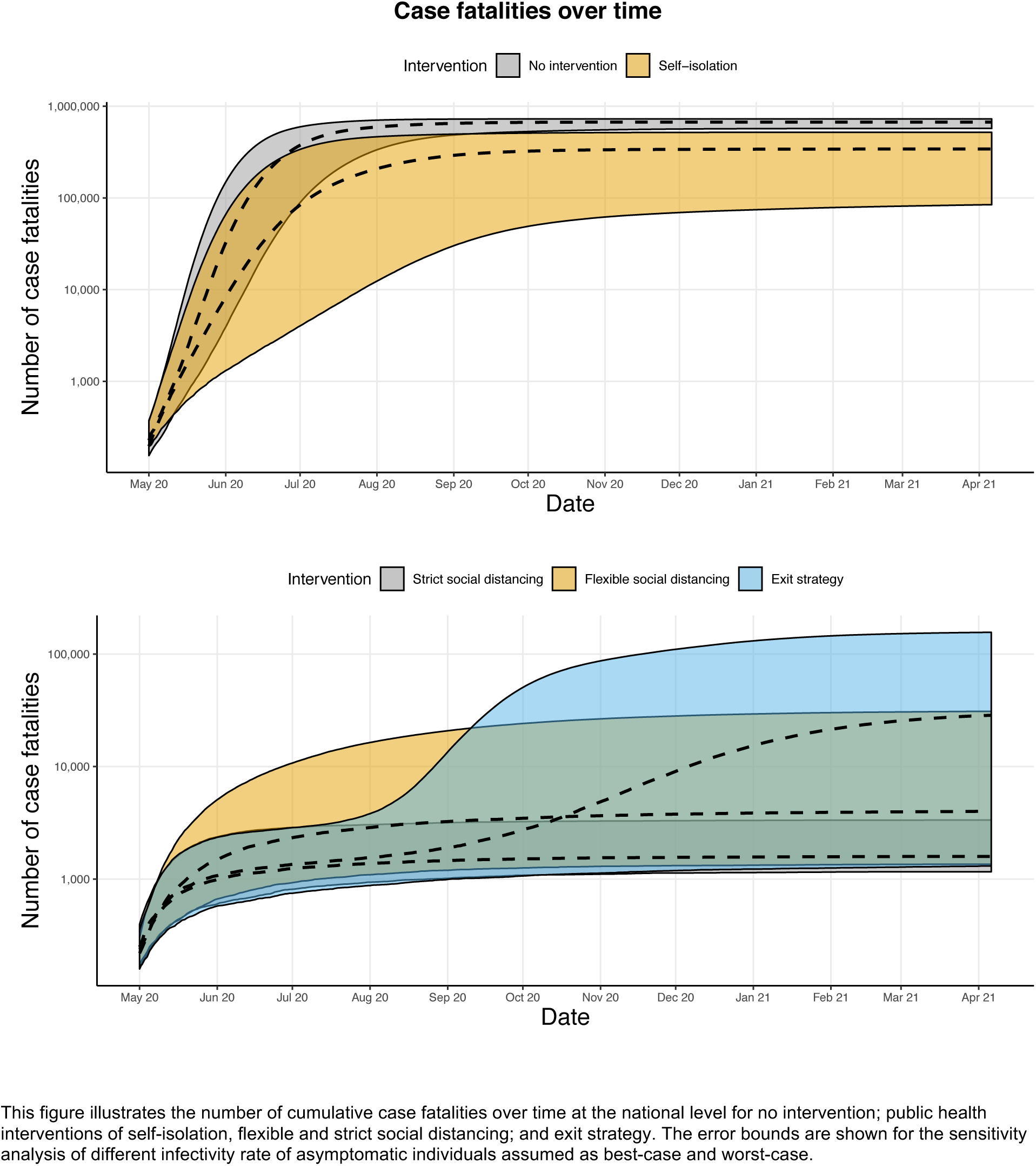
National-level estimates of cumulative case fatalities spread with sensitivity bands of infectivity of asymptomatic patients.

## Discussion

Our results, to no surprise, indicate that case fatalities and ICU hospitalizations for Pakistan will be high without any public health interventions. Our simulation showed that strict social distancing can potentially prevent a large number of deaths and ICU-hospitalizations. Our base case analysis showed total cumulative case fatalities of 1,588 deaths with the virus expected to peak in terms of the number of required ICU-hospitalizations at 266 persons by the end of May 2020. While the health benefits of strict social distancing will be high, the government faces an important dilemma of potentially severe economic downfall. Consideration of a temporary strict social distancing strategy with gradual return of the lower-risk Pakistani population, as simulated in our exit strategy scenario, may an effective compromise between public health and economy of Pakistani population.

Our model and analyses are not without limitations. COVID-19 is a new disease caused by a novel virus, and thus there are inevitable uncertainties in the data inputs that were utilized for this modelling study. In this regard, our prediction of the epidemic duration and size should be interpreted as applicable to the confirmed cases based on the testing method implemented in Pakistan. While we took into account the impact estimates of social distancing on regular activities from Google, the model also assumes homogenous mixing (contacts) between members of the population, without taking age group, fine-scale geography or other social factors into account. Additional work is required to add these aspects to such models. These figures largely represent people with smart phones and exclude the rural poor and slum dwellers. The network structure of each individual’s social circles also was not taken into consideration for this study, and there is also little information available on periodic clustering of people in mosques etc.

Regardless of these limitations, to our knowledge, this extended stochastic ICM model is the first to examine the effects of self-isolation and social distancing on the number of ICU hospitalizations required and case fatalities in South Asia. While there has been other modeling studies for COVID-19, these other studies have focused on regions such as Wuhan, China, and high-income countries such as Italy, Japan, South Korea, United States etc.^5,7,13,19,20^ This model may also be one of the first to take into account sub-national geographic factors, such as the population size and urban-to-rural distribution of different provinces of Pakistan, as well their differing hospital capacities.

The growing number of deaths and serious illnesses due to COVID-19 in Pakistan, and periodic suggestions of under-reporting of deaths^20^, suggest that we may be at a relatively early stage of the epidemic with an upsurge expected in the next few weeks as the circulation of the virus increases. The government is working to improve the number of critical care beds and ventilator capacity in public sector hospitals, but overall testing rates remain low and the rigidity of imposition of physical distancing measures and lock-downs, variable. Our model provides both short term projections as well as a trajectory in response to various non-pharmacological options. As Pakistan prepares for the long-haul of the COVID-19 pandemic, we plan to update the model with new outbreak data as they become available. Our mathematical simulation demonstrates the application of our model to the COVID-19 pandemic in Pakistan. Our model can fit the reported data well and predicts the appearance of an epidemic peak under different public health interventions, until the infection levels decrease and approach an endemic state in the long run. We plan to make our modelling approaches available for other investigators and countries to use beyond Pakistan.

There is much concern that preventive measures such as physical distancing, lock downs and curfews, being imposed as standard measures may not be readily applicable in countries with complex social conditions (such as conflict, migratory populations and large congested slums). Such circumstances require local adaptation of measures based on models that can be adapted to local realities and are flexible. We believe that our model allows that level of national and subnational planning and we intend to adapt it to the general situation in South Asia and also the local response in Pakistan.

## Data Availability

All of the data used for this study are made available in the manuscript

## Contributors

ZAB conceptualised and designed the study. ZAB, OH, JJHP, TC, KT, and EJM acquired, analysed, and interpreted data. ZAB, OH, and JJHP drafted the manuscript. All authors critically revised the manuscript for important intellectual content. OH and TC did the statistical analysis. ZAB obtained funding. ZAB provided administrative, technical, or material support, ZAB, KT, and EJM supervised the study.

## Acknowledgements

We have no acknowledgements to make.

